# Multiple Myeloma and SARS-CoV-2 Infection: Clinical Characteristics and Prognostic Factors of Inpatient Mortality

**DOI:** 10.1101/2020.06.29.20142455

**Authors:** Joaquín Martínez-López, María-Victoria Mateos, Cristina Encinas, Anna Sureda, José Ángel Hernández-Rivas, Ana López de la Guía, Diego Conde, Isabel Krsnik, Elena Prieto, Rosalía Riaza Grau, Mercedes Gironella, María Jesús Blanchard, Nerea Caminos, Carlos Fernández de Larrea, María Alicia Senin, Fernando Escalante, José Enrique de la Puerta, Eugenio Giménez, Pilar Martínez-Barranco, Juan José Mateos, Luis Felipe Casado, Joan Bladé, Juan José Lahuerta, Javier De la Cruz, Jesús San-Miguel

## Abstract

There is limited information on the characteristics, pre-admission prognostic factors, and outcomes of patients with multiple myeloma (MM) hospitalized with coronavirus disease 2019 (COVID-19). This retrospective case series investigated characteristics and outcomes of 167 MM patients hospitalized with COVID-19 reported from 73 hospitals within the Spanish Myeloma Collaborative Group network in Spain between March 1 and April 30, 2020. Outcomes were compared with a randomly selected contemporary cohort of 167 age-/sex-matched non-cancer patients with COVID-19 admitted at 6 participating hospitals. Common demographic, clinical, laboratory, treatment, and outcome variables were collected; specific disease status and treatment data were collected for MM patients. Among the MM and non-cancer patients, median age was 71 years and 57% of patients were male in each series, and 75% and 77% of patients, respectively, had at least one comorbidity. COVID-19 clinical severity was moderate-severe in 77% and 89% of patients and critical in 8% and 4%, respectively. Supplemental oxygen was required by 47% and 55% of MM and non-cancer patients, respectively, and 21%/9% vs 8%/6% required non-invasive/invasive ventilation. Inpatient mortality was 34% and 23% in MM and non-cancer patients, respectively. Among MM patients, inpatient mortality was 41% in males, 42% in patients aged >65 years, 49% in patients with active/progressive MM at hospitalization, and 59% in patients with comorbid renal disease at hospitalization, which were independent prognostic factors of inpatient mortality on adjusted multivariate analysis. This case series demonstrates the increased risk and identifies predictors of inpatient mortality among MM patients hospitalized with COVID-19.

**Key Points:**

- There is an increased risk of inpatient mortality (34% vs 23%) in MM vs age-/sex-matched non-cancer patients hospitalized with COVID-19.
- Adverse prognostic factors at admission for inpatient mortality in MM patients include age >65 y, male sex, renal disease, and active MM.

## INTRODUCTION

In December 2019, a cluster of patients with pneumonia linked to a previously unknown coronavirus, now named severe acute respiratory syndrome coronavirus 2 (SARS-CoV-2), was detected in Wuhan, China.^1^ This rapidly evolved into a pandemic of the heterogeneous clinical disease named coronavirus disease 2019 (COVID-19) that has challenged healthcare systems and led to thousands of deaths worldwide. The clinical presentation is highly variable, ranging from asymptomatic cases to severe respiratory infections. It is associated with a pro-inflammatory status, leading to cytokine release syndrome and hypercoagulation, with multi-organ damage and fatal outcome.^2^ It was soon recognized that various factors, including increased age (> 65 years, and particularly > 75 years), male sex, and presence of comorbidities (cardiovascular comorbidity, obesity, hypertension, and diabetes) contributed substantially to the severity of COVID-19. Investigations of novel strategies for the prevention and treatment of COVID-19 (including anti-malarials, anti-viral drugs, antibiotics, anti-interleukin-6/-1 therapies, and anticoagulants), together with appropriate intensive supportive care, have aimed to improve patient outcome.^3,4^ There are cumulative data indicating that patients with cancer may be at increased risk for more severe COVID-19 and associated complications, including those receiving or not receiving treatment within the month prior to infection,^5,6^ although other recent results suggest mortality may be primarily associated with age, male sex, and comorbidities.^7^

Multiple myeloma (MM) is a malignant proliferation of clonal B lymphoid cells at the last stage of maturation (plasma cells) that are responsible for secretion of immunoglobulins (Igs). Impairment of Ig production, with coexistence of a monoclonal component (M-component) plus immunosuppression of normal Igs, leads to severely impaired humoral immunity. Additionally, MM patients display dysfunctional cellular and innate immunity; this complex failure of immunosurveillance mechanisms makes MM patients highly susceptible to viral and bacterial infections.^8^ Some MM treatments may also positively or negatively impact the immune system. The most common side-effect of MM treatment is the induction of cytopenia, which is clearly associated with increased susceptibility to bacterial and viral infections. Chemotherapy, particularly high-dose melphalan, results in these side-effects. Corticosteroids have well-established immunosuppressive effects. Proteasome inhibitors induce T-cell dysfunction and are associated with an increased risk for varicella zoster virus reactivation.^9^ Immunomodulatory drugs may have a protective effect by increasing natural killer (NK) and T cell function but are also associated with cytopenia and an increased risk for venous thromboembolism.^10^ CD38 monoclonal antibodies reduce NK and immunosuppressive regulatory T cells and are associated with higher incidences of viral and bacterial infections.^8,11,12^

To date there is very limited information regarding characteristics and outcomes of MM patients with COVID-19, with only a single case report^13^ and a small UK series of 75 heterogeneous patients that did not report a non-MM reference group of patients with COVID-19.^14^ Several international and national guidelines have been reported,^15-19^ providing guidance to physicians on the management of patients with MM during the COVID-19 pandemic; however, these recommendations are based on consensus and general disease concepts, and lack specific data derived from infected patients. To our knowledge this is the first large case-series study to describe comprehensively the clinical characteristics of COVID-19 in hospitalized MM patients, compare outcomes with a non-cancer cohort of COVID-19 patients, and identify pre-admission prognostic factors of inpatient mortality.

## METHODS

This multicenter case-series analysis was performed at a national level by the Spanish Myeloma Collaborative Group (Programa Español de Tratamientos en Hematología^14,20^/Grupo Español de Mieloma^8^). The study was approved by the Institutional Review Board (IRB) of Hospital Universitario 12 de Octubre (n 20/208). This case-series study includes MM patients who were admitted to a participating hospital with a SARS-CoV-2 polymerase chain reaction (PCR)-positive test between March 1 and April 30, 2020. Additionally, a series of patients with no cancer history, matched by age and sex, were randomly selected from cohorts of COVID-19 patients admitted during the same time period at 6 participating hospitals.

Clinical specimens were obtained by nasopharyngeal swab collection in accordance with Spanish disease control and prevention guidelines. Samples were processed at local microbiology laboratories, and SARS-CoV-2 one-step real-time reverse transcriptase (rRT)-PCR diagnostic assay was performed.

A modified version of a questionnaire proposed by the International Myeloma Society was used for data extraction. A common set of demographic, clinical, laboratory, treatment, and outcome variables were collected for the MM and non-cancer cohorts of hospitalized COVID-19 patients. Specific disease status and treatment data were obtained for MM patients. Data extraction was performed locally by hematology departments in participating hospitals and centrally processed by the PETHEMA/GEM data monitoring unit. Hospital outcome status was defined as discharge from hospital or receiving ongoing care (survivor group) or death (non-survivor group).

### Statistical Analysis

Demographic, clinical, laboratory, and therapy covariates and hospital outcomes were summarized using descriptive statistics. Continuous variables were reported as median and interquartile range (IQR). Generic COVID-19 patient characteristics and specific MM features were analyzed according to the two hospital outcome categories. Crude and adjusted odds ratios (OR) with 95% confidence intervals (CI) were estimated with logistic regression analysis for a predefined set of well-established prognostic factors. Pre-admission characteristics, MM status, and comorbidities constituted the reference model and were used to adjust the association of relevant MM features with mortality. To limit model overfitting, the number of events per candidate variable was restricted to >10. Calibration and discrimination of the resulting models were assessed using the Hosmer-Lemeshow goodness-of-fit test and the c-statistic.^21^ We assessed internal validity with a bootstrapping procedure that used 500 replicate samples to gain insight into the stability of the reference model.^22^ Analyses were generated using SAS/STAT software, Version 9.4, SAS Institute Inc.

## RESULTS

Between March 1 and April 30, 2020, 216 patients with MM and SARS-Cov-2 infection detected by RT-PCR were reported from 73 healthcare centers. Thirty patients were excluded because of a diagnosis of a plasma cell disorder other than MM; 19 patients were excluded because they did not require hospital admission (**Figure 1**). As of May 20, 2020, 1 patient remained hospitalized and receiving ongoing care. The temporal and geographical distribution of reported cases matched the distribution of the epidemic in Spain; 114 (68%) were confirmed in the first 3 weeks of the lockdown period (March 15 to April 4). Eligible cases were notified from 29 of 50 provinces (58%), with 108 (65%) reported from two provinces with high population excess mortality.

**Figure 1.**
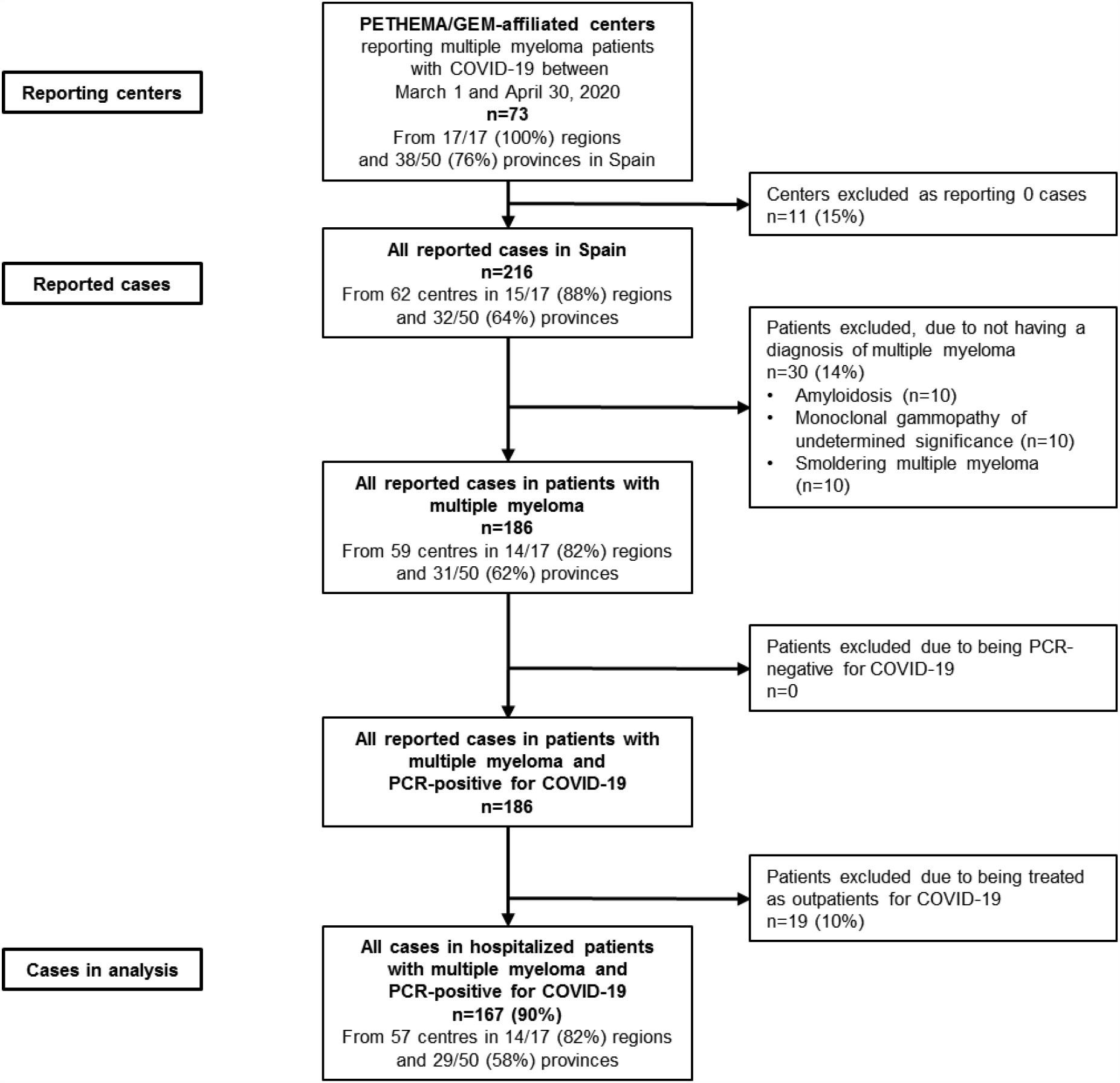
Flow diagram of patients with multiple myeloma admitted to hospital with COVID-19 who were included in the present analysis.

The characteristics and clinical features of the167 MM patients and the 167 non-cancer patients are described in **Table 1**. Concerning COVID-19 clinical features, 77% and 89% of MM and non-cancer patients, respectively, presented with moderate-severe disease, and 8% and 4% had critical disease. The proportion of patients requiring supplemental oxygen was 47% and 55% in MM and non-cancer patients, respectively. Numerically more MM patients required non-invasive and invasive ventilation (21% and 9%, respectively) compared to the non-cancer group (8% and 6%, respectively). At hospital admission, MM patients had numerically lower median levels of neutrophils, lymphocytes, and platelets, and numerically higher concentrations of D-dimers and ferritin compared with non-cancer patients (**Table 1**). Treatments administered to the two groups of patients are summarized in **Table 1**; 31% of MM patients received triple combination therapy including (hydroxy)chloroquine, azithromycin, and retrovirals compared with 42% of the non-cancer group. Regarding outcomes, inpatient mortality was 34% in the MM group and 23% in the non-cancer group.

**Table 1.** Pre-admission Features, COVID-19 Clinical, Laboratory, and Therapy Features and COVID-19 Outcome in Multiple Myeloma and Non-cancer Patients Hospitalized with COVID-19 Diagnosis.

|  | **Multiple myeloma patients (n=167), No. (%)** | **Non-cancer patients (n=167), No. (%)** |
| --- | --- | --- |
| **Preadmission features** |  | **Age-/sex-matched** |
| Age, median (IQR), y | 71 (62-78) | 71 (63-78) |
| Age groups |  |  |
| <65 y | 55 (33) | 55 (33) |
| 65-74 y | 55 (33) | 55 (33) |
| ≥75 y | 57 (34) | 57 (34) |
| Males | 95 (57) | 95 (57) |
| Females | 72 (43) | 72 (43) |
| Comorbidities |  |  |
| At least one | 126 (75) | 129 (77) |
| None | 41 (25) | 38 (23) |
| Cardiac disease | 35 (21) | 32 (19) |
| Pulmonary disease | 23 (14) | 34 (20) |
| Diabetes | 28 (17) | 36 (22) |
| Renal disease | 32 (19) | 13 (8) |
| Hypertension | 67 (40) | 79 (47) |
| **COVID-19 clinical features** |  |  |
| Clinical severity<sup>a</sup> |  |  |
| Mild | 26 (15) | 11 (7) |
| Moderate-severe | 128 (77) | 149 (89) |
| Critical | 13 (8) | 6 (4) |
| Respiratory support<sup>a</sup> |  |  |
| No supplemental oxygen | 39 (23) | 51 (31) |
| Supplemental oxygen | 78 (47) | 91 (55) |
| Non-invasive ventilation | 35 (21) | 14 (8) |
| Invasive ventilation | 15 (9) | 10 (6) |
| **COVID-19 laboratory measures at hospital admission, median (IQR)** |  |  |
| Absolute neutrophil count, $\times 10^9/L$ | 2.78 (1.7-4.7) | 4.8 (3.54-6.3) |
| Lymphocyte count, $\times 10^9/L$ | 0.515 (0.24-0.815) | 0.99 (0.7-1.42) |
| Platelets, $\times 10^9/L$ | 129 (78-176) | 215 (116-285) |
| D-dimers, $\mu g/mL$ | 0.861 (0.52-1.207) | 0.685 (0.422-1.383) |
| Ferritin, $ng/mL$ | 995 (486-2070) | 761 (401-1528) |
| **COVID-19 therapy** |  |  |
| (Hydroxy)chloroquine | 148 (89) | 157 (94) |
| Azithromycin | 91 (54) | 115 (69) |
| Antiretrovirals | 103 (62) | 103 (62) |
| Antiviral combination therapy |  |  |
| None | 11 (7) | 5 (3) |
| (Hydroxy)chloroquine only | 16 (10) | 14 (8) |
| (Hydroxy)chloroquine and retrovirals | 47 (28) | 31 (19) |
| (Hydroxy)chloroquine and azithromycin | 34 (20) | 42 (25) |
| (Hydroxy)chloroquine, azithromycin, and retrovirals | 51 (31) | 70 (42) |
| Other combination | 8 (5) | 5 (3) |
| Steroids | 83 (50) | 62 (37) |
| Anti-interleukin-6 receptor antibody therapy | 22 (13) | 27 (16) |
| Steroids and anti-interleukin-6 receptor antibody therapy | 21 (13) | 16 (10) |
| Heparin | 109 (65) | 117 (70) |
| **COVID-19 outcome (hospital outcome)** |  |  |
| Receiving ongoing care | 1 (<1) | 2 (1) |
| Hospital discharge | 110 (66) | 127 (76) |
| Hospital death | 56 (34) | 38 (23) |

<sup>a</sup>Data missing for 1 patient in the non-cancer group.
COVID-19, coronavirus disease 2019; IQR, interquartile range

### MM- and COVID-19-related characteristics according to hospital outcome

Pre-admission characteristics of MM patients according to hospital outcome are presented in **Table 2**; 56 patients (34%) died during hospitalization (non-survivor group), 110 (66%) were discharged, and 1 patient remained hospitalized receiving ongoing care (survivor group). Mortality was 41% in males and 24% in females. Mortality was 16% in patients aged < 65 years compared to 42% in those aged > 65 years, including 40% in patients aged 65–75 years and 44% in those aged > 75years. None of the female patients aged < 65 years died.

**Table 2.** Characteristics of Multiple Myeloma Patients with COVID-19, According to Hospital Outcome.

|  | Patients with hospital outcome, No. (%) |  |  |
| --- | --- | --- | --- |
|  | All (n=167) | Patients discharged (n=111)<sup>a</sup> | Patients who died (n=56) |
| Age, median (IQR), y | 71 (62-78) | 70 (58-77) | 75 (68-79) |
| Age groups |  | % shown as proportion of 'All' |  |
| ≤49 | 6 (4) | 6 (100) | 0 |
| 50-59 | 29 (17) | 24 (83) | 5 (17) |
| 60-69 | 41 (24) | 25 (61) | 16 (39) |
| 70-79 | 60 (36) | 37 (62) | 23 (38) |
| ≥80 | 31 (19) | 19 (61) | 12 (39) |
| Males | 95 (57) | 56 (59) | 39 (41) |
| Females | 72 (43) | 55 (76) | 17 (24) |
| **Multiple myeloma features** |  | % shown as proportion of 'All' |  |
| Monoclonal component |  |  |  |
| Immunoglobulin G | 83 (50) | 61 (73) | 22 (27) |
| Immunoglobulin A | 42 (25) | 25 (60) | 17 (40) |
| Immunoglobulin D | 2 (1) | 2 (100) | 0 (0) |
| Immunoglobulin M | 3 (2) | 2 (67) | 1 (33) |
| Non-secretory disease | 3 (2) | 2 (67) | 1 (33) |
| Light chain only disease | 34 (21) | 19 (56) | 15 (44) |
| International Staging System disease stage<sup>b</sup> |  |  |  |
| I | 47 (28) | 34 (72) | 13 (28) |
| II | 61 (37) | 42 (69) | 19 (31) |
| III | 57 (35) | 35 (61) | 22 (39) |
| Cytogenetic classification by FISH<sup>c</sup> |  |  |  |
| High risk | 36 (24) | 23 (64) | 13 (36) |
| Other/Standard risk | 114 (76) | 75 (66) | 39 (34) |
| Bone disease at diagnosis of multiple myeloma |  |  |  |
| Yes | 129 (77) | 86 (67) | 43 (33) |
| No | 38 (23) | 25 (66) | 13 (34) |
| Renal disease at diagnosis of multiple myeloma |  |  |  |
| Yes | 45 (27) | 22 (49) | 23 (51) |
| No | 122 (73) | 89 (73) | 33 (27) |
| Year of diagnosis of multiple myeloma (time since diagnosis), y |  |  |  |
| 2020 (0-3 m) | 25 (15) | 13 (52) | 12 (48) |
| 2019 ( 4-18 m) | 30 (18) | 24 (80) | 6 (20) |
| ≤2018 (≥18 m) | 112 (67) | 74 (66) | 38 (34) |
| Prior stem cell transplantation<sup>d</sup> |  |  |  |
| Yes | 51 (31) | 42 (82) | 9 (18) |
| No | 113 (69) | 67 (59) | 46 (41) |
| Prior lines of treatment for multiple myeloma<sup>e</sup> |  |  |  |
| 1 | 82 (51) | 58 (71) | 24 (29) |
| 2 | 39 (24) | 23 (59) | 16 (41) |
| >3 | 41 (25) | 27 (66) | 14 (34) |
| Prior therapy class |  |  |  |
| Proteasome inhibitor | 138 (83) | 92 (67) | 46 (33) |
| No proteasome inhibitor | 29 (17) | 19 (66) | 10 (34) |
| Immunomodulatory drug | 119 (71) | 80 (67) | 39 (33) |
| No immunomodulatory drug | 48 (29) | 31 (65) | 17 (35) |
| Monoclonal antibody | 38 (23) | 25 (66) | 13 (34) |
| No monoclonal antibody | 129 (77) | 86 (67) | 43 (33) |
| **Multiple myeloma status and comorbidities at hospital admission for COVID-19** |  | % shown as proportion of 'All' |  |
| Myeloma status |  |  |  |
| Active disease or progression | 43 (26) | 22 (51) | 21 (49) |
| Partial response | 72 (43) | 52 (72) | 20 (28) |
| Complete response | 52 (31) | 37 (71) | 15 (29) |
| Comorbidities |  |  |  |
| At least one | 126 (75) | 79 (63) | 47 (37) |
| None | 41 (25) | 32 (78) | 9 (22) |
| Cardiac disease | 35 (21) | 22 (63) | 13 (37) |
| No cardiac disease | 132 (79) | 89 (67) | 43 (33) |
| Pulmonary disease | 23 (14) | 14 (61) | 9 (39) |
| No pulmonary disease | 144 (86) | 97 (67) | 47 (33) |
| Diabetes | 28 (17) | 18 (64) | 10 (36) |
| No diabetes | 139 (83) | 93 (67) | 46 (33) |
| Renal disease | 32 (19) | 13 (41) | 19 (59) |
| No renal disease | 135 (81) | 9 (73) | 37 (27) |
| Hypertension | 67 (40) | 39 (58) | 28 (42) |
| No hypertension | 100 (60) | 71 (72) | 28 (28) |
| Immunoparesis<sup>f</sup> | 104 (69) | 67 (65) | 37 (31) |
| No immunoparesis | 46 (31) | 30 (65) | 16 (35) |

COVID-19, coronavirus disease 2019; FISH, fluorescence *in situ* hybridization; IQR, interquartile range
<sup>a</sup>Includes 1 patient who remains hospitalized receiving ongoing care. <sup>b</sup>Data missing for 2 patients who died in hospital. <sup>c</sup>Data missing for 17 patients, 13 who were discharged and 4 who died in hospital. <sup>d</sup>Data missing for 3 patients, 2 who were discharged and 1 who died in hospital. <sup>e</sup>Data missing for 5 patients, 3 who were discharged and 2 who died in hospital. <sup>f</sup>Data missing for 16 patients, 13 who were discharged and 3 who died in hospital.

Concerning MM features, inpatient mortality was 27% and 28% in patients with an IgG M-component and stage I disease at diagnosis, respectively. Cytogenetic abnormalities and presence of bone disease did not impact inpatient mortality. However, in patients with renal impairment at MM diagnosis inpatient mortality was 51% vs 27% in patients with normal renal function. Prior treatment with immunomodulatory drugs, proteasome inhibitors, or monoclonal antibodies did not impact inpatient mortality. (**Table 2**).

With regards to year of MM diagnosis, 12 of 25 patients (48%) diagnosed between January and April 2020 (during the emerging pandemic) did not survive COVID-19. To understand this high mortality rate we analyzed the characteristics of the cohort. Of note, 10 (83%) of the 12 non-survivors were male, 11 (92%) had a M-component other than IgG, 9 (75%) had International Staging System stage III disease, 7 (58%) presented with renal impairment, and 7 (58%) had active disease. By contrast, 82% of patients who had received autologous stem cell transplantation (ASCT) survived COVID-19 while 41% of the non-ASCT group died (**Table 2**).

Regarding MM status and comorbidities at the time of COVID-19 admission, in patients with active/progressive disease the inpatient mortality rate was 49% compared to 28% and 29% for patients in partial or complete response, respectively (**Table 2**). Patients with at least one comorbidity had an inpatient mortality rate of 37% vs 22% in those without comorbidities. The mortality rate was numerically higher in MM patients with vs without cardiac (37% vs 33%) or pulmonary (39% vs 33%) comorbidities or hypertension (42% vs 28%); presence of renal disease was associated with the numerically highest mortality rate (59%). Immunoparesis did not affect mortality rate.

**Table 3** summarizes clinical and laboratory features at admission according to hospital outcome. Patients with higher values (above the median) for neutrophil count, D-dimers, and ferritin had higher mortality rates (43%, 40%, and 32%, respectively) than those in patients with lower values (below the median; 25%, 22%, and 22%, respectively). The opposite effect was observed for lymphocyte and platelet counts, with lower values associated with higher mortality rate (40% vs 28% and 41% vs 25%, respectively).

**Table 3.** Patient Characteristics at Hospital Admission for COVID-19, According to Hospital Outcome.

|  | Patients with hospital outcome, No. (%) |  |  |
| --- | --- | --- | --- |
|  | All (n=167) | Patients discharged (n=111)<sup>a</sup> | Patients who died (n=56) |
| **Clinical features during admission** |  | % shown as proportion of 'All' |  |
| Clinical severity |  |  |  |
| Mild | 26 (16) | 24 (92) | 2 (8) |
| Moderate-severe | 128 (77) | 85 (66) | 43 (34) |
| Critical | 13 (8) | 2 (15) | 11 (85) |
| Respiratory support |  |  |  |
| No supplemental oxygen | 39 (23) | 36 (92) | 3 (8) |
| Supplemental oxygen | 78 (47) | 56 (72) | 22 (28) |
| Non-invasive ventilation | 35 (21) | 13 (37) | 22 (63) |
| Invasive ventilation | 15 (9) | 6 (40) | 9 (60) |
| **Laboratory measures at hospital admission** |  | % shown as proportion of 'All' |  |
| Absolute neutrophil count<sup>b</sup> |  |  |  |
| Above median | 80 (50) | 46 (57) | 34 (43) |
| Below median | 79 (50) | 59 (75) | 20 (25) |
| Lymphocyte count<sup>c</sup> |  |  |  |
| Above median | 79 (50) | 57 (72) | 22 (28) |
| Below median | 78 (50) | 47 (60) | 31 (40) |
| Platelets<sup>d</sup> |  |  |  |
| Above median | 77 (51) | 58 (75) | 19 (25) |
| Below median | 75 (49) | 44 (59) | 31 (41) |
| D-dimers<sup>e</sup> |  |  |  |
| Above median | 67 (51) | 40 (60) | 27 (40) |
| Below median | 64 (49) | 50 (78) | 14 (22) |
| Ferritin<sup>f</sup> |  |  |  |
| Above median | 41 (50) | 28 (68) | 13 (32) |
| Below median | 41 (50) | 32 (78) | 9 (22) |
| **COVID-19 therapy** |  | % shown as proportion of 'All' |  |
| (Hydroxy)chloroquine | 148 (89) | 101 (68) | 47 (32) |
| No (hydroxy)chloroquine | 19 (11) | 10 (53) | 9 (47) |
| Azithromycin | 91 (54) | 62 (68) | 29 (32) |
| No azithromycin | 76 (46) | 49 (64) | 27 (36) |
| Antiretrovirals | 103 (62) | 65 (63) | 38 (37) |
| No antiretrovirals | 64 (38) | 46 (72) | 18 (28) |
| Antiviral combination therapy |  |  |  |
| None | 11 (6) | 6 (55) | 5 (45) |
| (Hydroxy)chloroquine only | 16 (10) | 11 (69) | 5 (31) |
| (Hydroxy)chloroquine and retrovirals | 47 (28) | 30 (64) | 17 (36) |
| (Hydroxy)chloroquine and azithromycin | 34 (20) | 27 (79) | 7 (21) |
| (Hydroxy)chloroquine, azithromycin, and retrovirals | 51 (31) | 33 (65) | 18 (35) |
| Other combination therapy | 8 (5) | 4 (50) | 4 (50) |
| Steroids | 83 (50) | 51 (61) | 32 (39) |
| No steroids | 84 (50) | 60 (71) | 24 (29) |
| Anti-interleukin-6 receptor antibody therapy | 22 (13) | 15 (68) | 7 (32) |
| No anti-interleukin-6 receptor antibody therapy | 145 (87) | 96 (66) | 49 (34) |
| Steroids and anti-interleukin-6 receptor antibody therapy<sup>f</sup> | 21 (13) | 14 (67) | 7 (33) |
| No steroids and anti-interleukin-6 receptor antibody therapy<sup>f</sup> | 143 (87) | 95 (66) | 48 (34) |
| Heparin | 109 (65) | 77 (71) | 32 (29) |
| No heparin | 58 (35) | 34 (59) | 24 (41) |

COVID-19, coronavirus disease 2019; IQR, interquartile range
<sup>a</sup>Includes 1 patient who remains hospitalized receiving ongoing care. <sup>b</sup>Data missing for 8 patients, 6 who were discharged and 2 who died in hospital. <sup>c</sup>Data missing for 11 patients, 8 who were discharged and 3 who died in hospital. <sup>d</sup>Data missing for 16 patients, 10 who were discharged and 6 who died in hospital. <sup>e</sup>Data missing for 36 patients, 21 who were discharged and 15 who died in hospital. <sup>f</sup>Data missing for 85 patients, 51 who were discharged and 34 who died in hospital. <sup>g</sup>Data missing for 3 patients, 2 who were discharged and 1 who died in hospital.

The association with inpatient mortality was estimated for 9 prognostic factors assessed before admission (**Table 4**). In an unadjusted analysis, 7 factors were associated with increased odds of death: male sex, age > 65 years, renal disease and hypertension as comorbidities, diagnosis of MM in 2020, renal disease at diagnosis, and active/progressive disease at the time of COVID-19 hospitalization (Table 4). IgG M-component and prior ASCT were associated with lower inpatient mortality. In an adjusted analysis, male sex, age > 65 years, active/progressive disease, and renal disease as a comorbidity were independent prognostic factors of inpatient mortality. The combination of this set of independent factors constituted a stable prognostic model of inpatient mortality; the bootstrapping procedure corrected the discriminatory ability of this reference model in terms of *c*-statistic from 0.79 to 0.765. No other candidate variable improved the prognostic performance of the reference model in terms of calibration and discrimination (**Table 4**).

**Table 4.** Prognostic Factors of Inpatient Mortality in Multiple Myeloma Patients Hospitalized with COVID-19.

| Prognostic Factors<sup>a</sup> | N | Inpatient Mortality,<br>No. (%) | Unadjusted Analysis<sup>b</sup> |  | Adjusted Analysis<sup>c</sup> |  | GoF | c-statistic |
| --- | --- | --- | --- | --- | --- | --- | --- | --- |
|  |  |  | Odds Ratio<br>(95% CI) | P value | Odds Ratio<br>(95% CI) | P value | P value | (95% CI) |
| All patients | 167 | 56 (34) |  |  |  |  |  |  |
| Age >65 y | 112 | 47 (42) | 3.7 (1.6-8.3) | 0.002 | 3. (1.4-8.4) | 0.006 | Na | Na |
| Males | 95 | 39 (41) | 2.3 (1.1-4.4) | 0.018 | 3.8 (1.7-8.4) | 0.001 | Na | Na |
| Multiple myeloma comorbidities and status at hospital admission for COVID-19 |  |  |  |  |  |  |  |  |
| Renal disease | 32 | 19 (59) | 3.9 (1.7-8.7) | <0.001 | 4.6 (1.9-11.3) | <0.001 | Na | Na |
| Hypertension | 67 | 28 (42) | 1.8 (0.96-3.5) | 0.065 | 1.7 (0.8-3.5) | 0.18 | Na | Na |
| Active disease or progression | 43 | 21 (49) | 2.4 (1.2-4.9) | 0.015 | 2.7 (1.2-6.0) | 0.017 | Na | Na |
| Reference model: |  |  |  |  |  |  | 0.7 | 0.79 (0.72-0.86) |
| Multiple myeloma features at diagnosis and treatment |  |  |  |  |  |  |  |  |
| Monoclonal component, immunoglobulin G | 83 | 22 (27) | 0.5 (0.3-1.0) | 0.05 | 0.6 (0.3-1.3) | 0.18 | 0.6 | 0.80 (0.73-0.87) |
| Renal disease at diagnosis | 45 | 23 (51) | 2.8 (1.4-5.7) | 0.004 | 1.3 (0.4-3.7) | 0.6 | 0.8 | 0.79 (0.72-0.86) |
| Diagnosis in 2020 (time since diagnosis $\leq 3$ m) | 25 | 12 (48) | 2.0 (0.9-4.9) | 0.10 | 2.7 (0.7-5.8) | 0.19 | 0.5 | 0.79 (0.72-0.86) |
| Prior stem cell transplantation | 51 | 9 (17) | 0.3 (0.1-0.7) | 0.004 | 0.6 (0.2-1.7) | 0.4 | 0.4 | 0.79 (0.71-0.86) |

<sup>a</sup>Predefined set of well-established prognostic factors assessed before admission; all variables were dichotomized according to standard categories.
<sup>b</sup>Crude odds ratio and 95% confidence interval.
<sup>c</sup>Each logistic model included age, sex, myeloma status, comorbidities (hypertension, renal disease) at diagnosis of COVID-19 [reference model], and one variable from the 'Multiple myeloma at diagnosis and treatment' set. Calibration and discrimination of the models were assessed with the Hosmer-Lemeshow goodness-of-fit test (GoF *P* value) and the c-statistic.
CI, confidence interval; COVID-19, coronavirus disease 2019; GoF, goodness-of-fit; Na, not applicable

## DISCUSSION

This study provides a comprehensive description of the characteristics and outcome of MM patients hospitalized with confirmed SARS-CoV-2 infection in Spain. Mortality was 50% higher in MM patients (34%) vs non-cancer patients (23%). The main predictors of inpatient mortality for MM were male sex, age > 65 years, renal disease, and active/progressive disease.

This is a highly representative series for gaining insights into the impact of COVID-19 in the MM population; all Spanish hospitals were put under the governance of the Ministry of Health, and the structure of PETHEMA/GEM includes most centers treating MM patients within Spanish territory. The selected time-frame corresponded to peak COVID-19 incidence and population excess mortality in Spain. Additionally, to gain a more precise picture of the differential features of these patients, we obtained clinical and laboratory characteristics and hospital outcomes from a concomitant series of SARS-CoV-2-infected non-cancer patients. To assure uniformity, the study focused solely on patients requiring hospital admission – we did not include MM patients who were treated as outpatients for COVID-19 and either were PCR-positive or just had COVID-19-compatible symptomatology since, in such a population, obtaining a complete case history could not be guaranteed.

Participating centers covered the whole geographical territory of Spain, although half of the cases were reported from the Madrid region, where a higher number of COVID-19 cases were confirmed at the national level. The large number of participating hospitals (n = 73) ensured a representative mix of cases. The central data monitoring unit worked closely with local sites to confirm data and on patient follow-up. Participating centers were long-term contributors to the PETHEMA/GEM research program; local data extractors at each site implemented common data element definitions.

The selection of prognostic factors for estimating adjusted associations with inpatient mortality rate was based on clinical credibility and restricted in number to limit model overfitting. The assessment of internal validity confirmed the stability of the model and that factor estimates were realistic for similar future patients. We assessed the risk of study bias with the QIPS tool for prognostic factors studies and concluded that the reported results are unlikely to be distorted, spurious, or biased.^23^ The independent prognostic factors that we identified were measurements available prior to hospital admission. A prognostic model including these factors could be developed to risk-stratify MM patients and to implement preventive strategies in the event of a healthcare crisis.

As noted, inpatient mortality was higher in MM vs non-cancer patients. This occurred despite the similarity between groups in the clinical severity of COVID-19. Rates of comorbidities appeared comparable between groups, except for renal disease, which has emerged as a critical feature in the outcome of SARS-CoV-2-infected MM patients. Interestingly, presence of lymphopenia and higher levels of D-dimers and ferritin, laboratory findings associated with higher mortality rates in several global COVID-19 patient series,^2,4,24,25^ were more common in MM vs non-cancer patients. Nevertheless, the mortality rate observed for MM patients is similar to that reported in patients with other cancers (28.6%) and hematologic diseases (33%), as well as immunocompromised patients (27.8%).^5,7,26,27^

The higher inpatient mortality rate in MM vs non-cancer patients can be explained through patient-related factors and MM features, besides the biology of the disease itself. Regarding patient-related factors, age > 65 years and male sex emerged as two of the most important risk factors predicting inpatient mortality. Notably, and in contrast to non-cancer COVID-19 populations, in which 75 years has been reported as the cutoff predicting peak risk of death, the mortality rate in MM did not differ above 65 years of age, suggesting that there are other factors intrinsic to MM that influence outcome. Regarding MM features, renal insufficiency is a major hallmark of MM (one of the four myeloma-defining events); in the present series its presence at hospital admission for COVID-19 emerged as the most important factor for survival (OR = 3.8). Renal impairment at diagnosis was also an adverse factor, but this significance was lost in the adjusted multivariate analysis. However, the strength of association with inpatient mortality of renal insufficiency at admission increased and independent prognostic value was retained in the multivariate analysis. Indeed, kidney disease as a prognostic factor predicting increased inpatient mortality rate in a general patient population has been previously described in a single-center study in China.^28^ By contrast, hypertension as a comorbidity was not a significant factor in the adjusted model. Other MM features such as high-risk cytogenetics and immunoparesis did not influence outcome; the latter is of interest since humoral immunodeficiency, which is common in MM, would be considered a potential factor for elevated risk. Additionally, we observed that IgG isotype and stage I disease were associated with lower mortality. Regarding MM status at hospital admission for COVID-19, as would be expected the presence of uncontrolled disease had a detrimental effect on survival (OR = 2.6) and was one of the independent factors predicting outcome. This would help explain the significantly higher mortality rate observed in patients diagnosed at the time of pandemic (January-April 2020), since in most their disease was not adequately controlled; it is well known that newly diagnosed patients have an elevated risk of mortality due to infectious complications in the first 3 months of treatment. The benefit of disease control (complete/partial response) in the context of COVID-19 would likewise explain the favorable outcome observed in patients who had previously received ASCT, since most of them were in response.

The non-participation of MM patients with unconfirmed COVID-19 or who were not hospitalized is a limitation in terms of the broader applicability of the study results. Further studies could be conducted to implement our analysis protocol in a less restricted population of patients. Another limitation is that our study is not able to identify optimal management and treatment of newly diagnosed MM patients with COVID-19.

In conclusion, MM patients hospitalized with SARS-CoV-2 infection have a higher inpatient mortality rate than non-cancer patients. This had been suspected since the beginning of the COVID-19 pandemic, and consequently national and international societies have published or made general recommendations about optimal management of MM patients. Our study assessed the impact of prior MM treatments based on immunomodulatory drugs, proteasome inhibitors, or monoclonal antibodies, and none of these influenced in the inpatient mortality rate. Although we cannot answer the question of whether to treat or not – and how – during the pandemic, we can stress the need for appropriate disease control in all patients, to minimize hospital visits, particularly in the most vulnerable MM populations such as male patients, those aged >65 years (who are already at high risk, even at this relatively young age), and those with renal impairment, which is the most critical factor. Accordingly, MM patients requiring treatment due to active disease should be tested for COVID-19 by PCR and, if negative, we should proceed to optimize treatment in the most protective environment possible. In patients with non-active disease, MM treatment could be postponed for 1-2 months; however, if patients are receiving maintenance therapy, treatment should continue. Further research is required to identify the most effective treatments for COVID-19, as well as vaccination strategies that should be employed as early as possible in the most vulnerable patients. Future studies should validate criteria for stratifying MM patients who are at greater risk in a healthcare crisis such as the COVID-19 pandemic and who could benefit from reducing their risk-exposure through a more stringent preventive strategy.

## Data Availability

The data that support the findings of this study are available from the corresponding author, upon reasonable request

## ACKNOWLEDGMENTS

We would like to thank Roberto Maldonado from the PETHEMA Foundation for data monitoring and technical support, and other members of the Grupo Español de Mieloma (GEM) participating in the study:

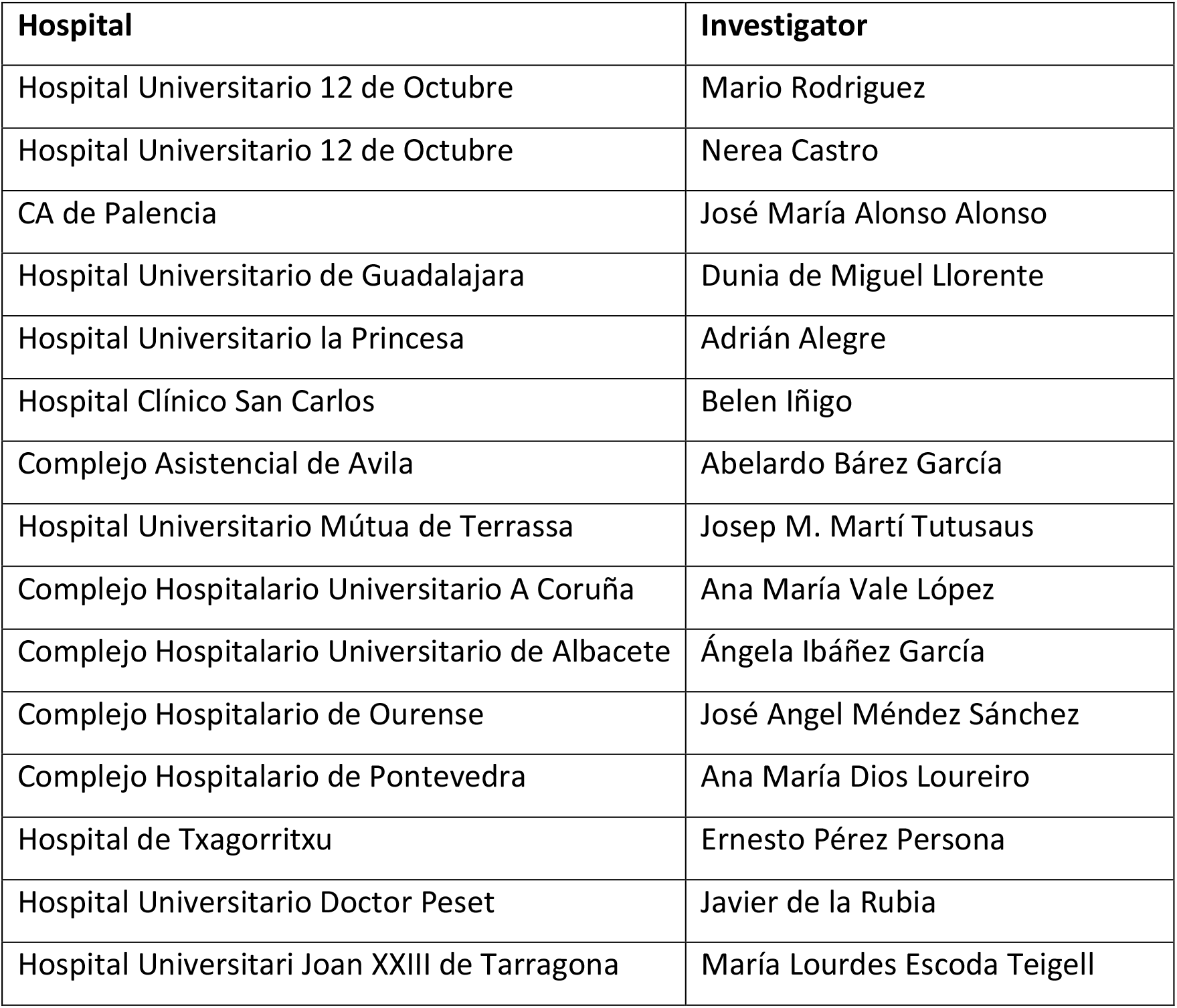
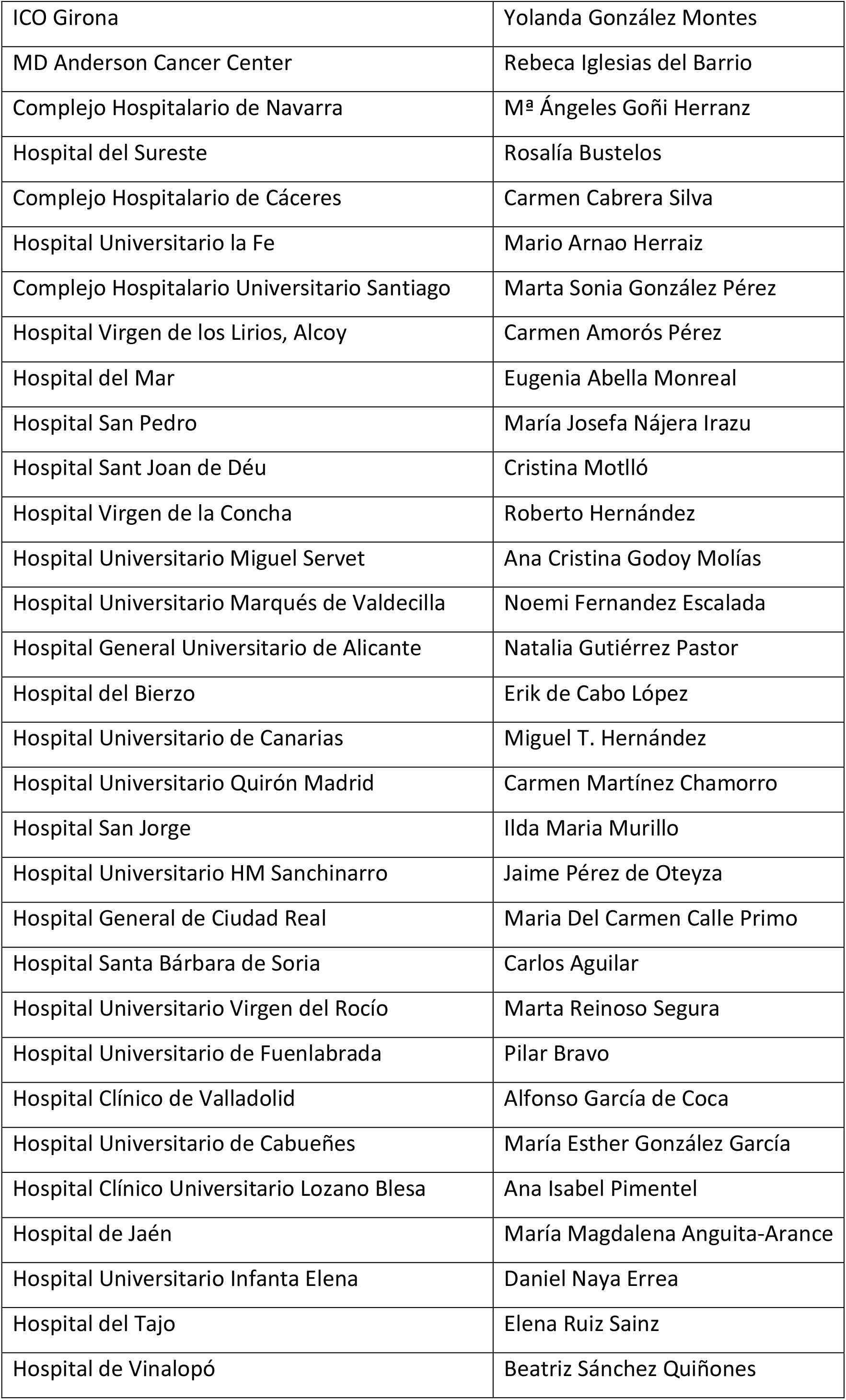
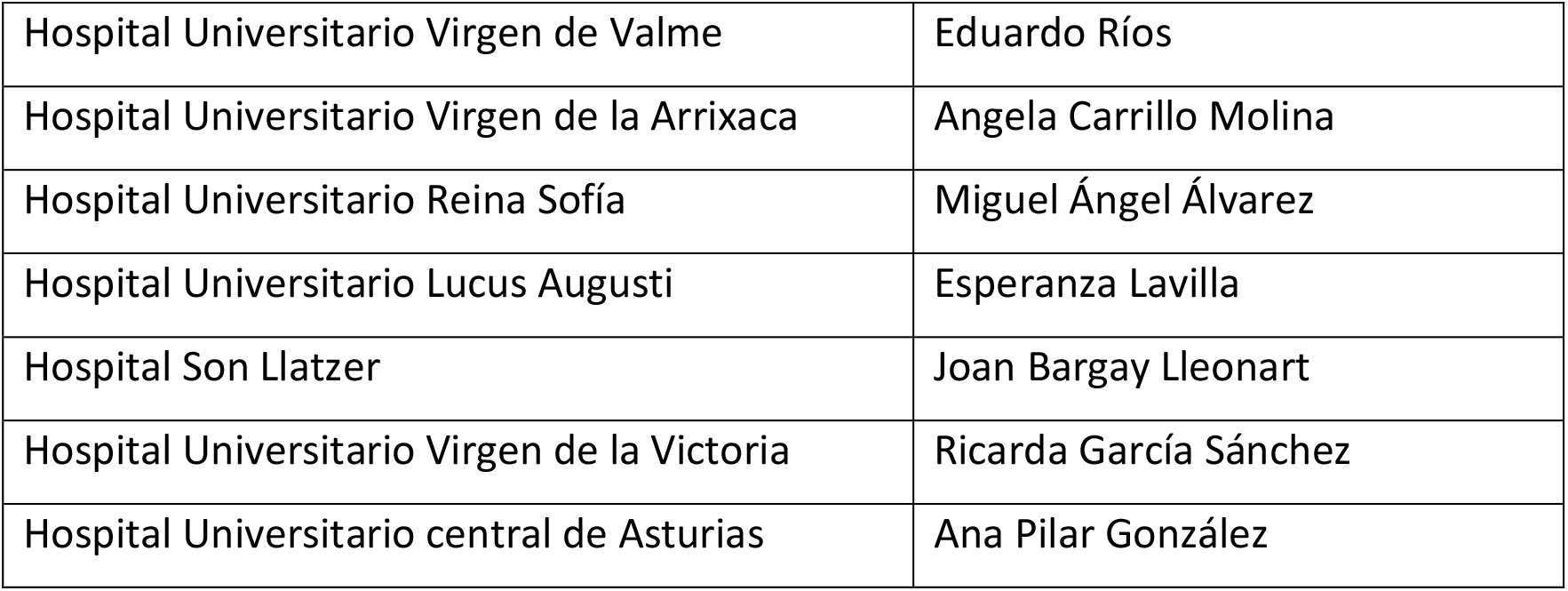

| Hospital | Investigator |
| --- | --- |
| Hospital Universitario 12 de Octubre | Mario Rodriguez |
| Hospital Universitario 12 de Octubre | Nerea Castro |
| CA de Palencia | José María Alonso Alonso |
| Hospital Universitario de Guadalajara | Dunia de Miguel Llorente |
| Hospital Universitario la Princesa | Adrián Alegre |
| Hospital Clínico San Carlos | Belen Iñigo |
| Complejo Asistencial de Avila | Abelardo Báñez García |
| Hospital Universitario Mútua de Terrassa | Josep M. Martí Tutusaus |
| Complejo Hospitalario Universitario A Coruña | Ana María Vale López |
| Complejo Hospitalario Universitario de Albacete | Ángela Ibáñez García |
| Complejo Hospitalario de Ourense | José Angel Méndez Sánchez |
| Complejo Hospitalario de Pontevedra | Ana María Dios Loureiro |
| Hospital de Txagorritxu | Ernesto Pérez Persona |
| Hospital Universitario Doctor Peset | Javier de la Rubia |
| Hospital Universitari Joan XXIII de Tarragona | María Lourdes Escoda Teigell |
| ICO Girona | Yolanda González Montes |
| MD Anderson Cancer Center | Rebeca Iglesias del Barrio |
| Complejo Hospitalario de Navarra | M<sup>a</sup> Ángeles Goñi Herranz |
| Hospital del Sureste | Rosalía Bustelos |
| Complejo Hospitalario de Cáceres | Carmen Cabrera Silva |
| Hospital Universitario la Fe | Mario Arnao Herraiz |
| Complejo Hospitalario Universitario Santiago | Marta Sonia González Pérez |
| Hospital Virgen de los Lirios, Alcoy | Carmen Amorós Pérez |
| Hospital del Mar | Eugenia Abella Monreal |
| Hospital San Pedro | María Josefa Nájera Irazu |
| Hospital Sant Joan de Déu | Cristina Motlló |
| Hospital Virgen de la Concha | Roberto Hernández |
| Hospital Universitario Miguel Servet | Ana Cristina Godoy Molías |
| Hospital Universitario Marqués de Valdecilla | Noemi Fernandez Escalada |
| Hospital General Universitario de Alicante | Natalia Gutiérrez Pastor |
| Hospital del Bierzo | Erik de Cabo López |
| Hospital Universitario de Canarias | Miguel T. Hernández |
| Hospital Universitario Quirón Madrid | Carmen Martínez Chamorro |
| Hospital San Jorge | Ilda Maria Murillo |
| Hospital Universitario HM Sanchinarro | Jaime Pérez de Oteyza |
| Hospital General de Ciudad Real | Maria Del Carmen Calle Primo |
| Hospital Santa Bárbara de Soria | Carlos Aguilar |
| Hospital Universitario Virgen del Rocío | Marta Reinoso Segura |
| Hospital Universitario de Fuenlabrada | Pilar Bravo |
| Hospital Clínico de Valladolid | Alfonso García de Coca |
| Hospital Universitario de Cabueñes | María Esther González García |
| Hospital Clínico Universitario Lozano Blesa | Ana Isabel Pimentel |
| Hospital de Jaén | María Magdalena Anguita-Arance |
| Hospital Universitario Infanta Elena | Daniel Naya Errea |
| Hospital del Tajo | Elena Ruiz Sainz |
| Hospital de Vinalopó | Beatriz Sánchez Quiñones |
| Hospital Universitario Virgen de Valme | Eduardo Ríos |
| Hospital Universitario Virgen de la Arrixaca | Angela Carrillo Molina |
| Hospital Universitario Reina Sofía | Miguel Ángel Álvarez |
| Hospital Universitario Lucus Augusti | Esperanza Lavilla |
| Hospital Son Llatzer | Joan Bargay Lleonart |
| Hospital Universitario Virgen de la Victoria | Ricarda García Sánchez |
| Hospital Universitario central de Asturias | Ana Pilar González |

We thank Steve Hill, PhD, of FireKite, an Ashfield company, part of UDG Healthcare plc, for editorial assistance in the preparation of this manuscript for submission, which was funded by PETHEMA/GEM.

## AUTHORSHIP CONTRIBUTIONS

Drs Martinez-Lopez, Mateos, De la Cruz, and San-Miguel had full access to all of the data in the study and take responsibility for the integrity of the data and the accuracy of the data analysis.

### Concept and design

Martinez-Lopez, Mateos, De la Cruz, and San-Miguel.

### Acquisition, analysis, or interpretation of data

Martinez-Lopez, Mateos, Encinas, Sureda, Hernández-Rivas, López de la Guía, Conde, Krsnik, Prieto, Riaza-Grau, Gironella, Blanchard, Caminos, Fernández de Larrea, Senin, Escalante, de la Puerta, Giménez, Martínez-Barranco, Mateos, Casado, Bladé, Lahuerta, De la Cruz, San-Miguel.

### Drafting of the manuscript

Martinez-Lopez, Mateos, De la Cruz, and San-Miguel.

### Critical revision of the manuscript for important intellectual content

Martinez-Lopez, Mateos, Encinas, Sureda, Hernández-Rivas, López de la Guía, Conde, Krsnik, Prieto, Riaza-Grau, Gironella, Blanchard, Caminos, Fernández de Larrea, Senin, Escalante, de la Puerta, Giménez, Martínez-Barranco, Mateos, Casado, Bladé, Lahuerta, De la Cruz, and San-Miguel.

### Statistical analysis

De la Cruz.

